# Clinicopathological and prognostic correlations of plasma biomarkers in neurodegenerative diseases

**DOI:** 10.64898/2025.12.07.25341788

**Authors:** Maura Malpetti, Leonidas Chouliaras, Alexander G Murley, Peter Swann, Ian Coyle-Gilchrist, W Richard Bevan-Jones, George Savulich, Marlou N Perquin, Julie Wiggins, Lucy Bowns, Li Su, Laura Hughes, Annelies Quaegebeur, Kieren Allinson, Timothy Rittman, Young Hong, Tim Fryer, Rhys Roberts, Nicholas J Ashton, Kaj Blennow, Henrik Zetterberg, John T O’Brien, James B Rowe

**Affiliations:** University of Cambridge Department of Clinical Neurosciences and Cambridge University Hospitals NHS Trust, Cambridge, CB2 0SZ, UK; UK Dementia Research Institute at University of Cambridge, Cambridge CB2 0XY, UK; Department of Psychiatry, University of Cambridge, Cambridge, CB2 0QQ, UK; Essex Partnership University NHS Foundation Trust, St Margaret’s Hospital Epping, CM16 6TN5; Sheffield Institute of Translational Neuroscience, University of Sheffield, S10 2HQ, UK; Department of Psychiatry and Neurochemistry, Institute of Neuroscience and Physiology, The Sahlgrenska Academy, University of Gothenburg, Mölndal, S-431 80, Sweden; Banner Alzheimer’s Institute and University of Arizona, Phoenix, AZ, USA; Banner Sun Health Research Institute, Sun City, AZ 85351, USA; Clinical Neurochemistry Laboratory, Sahlgrenska University Hospital, Mölndal, S-431 80, Sweden; UK Dementia Research Institute at UCL, London, WC1N 6BG, UK; Department of Neurodegenerative Disease, UCL Institute of Neurology, London, WC1N 6BG, UK; Hong Kong Center for Neurodegenerative Diseases, Clear Water Bay, Hong Kong, China; Wisconsin Alzheimer’s Disease Research Center, University of Wisconsin School of Medicine and Public Health, University of Wisconsin-Madison, Madison, WI, 53792, USA; Centre for Brain Research, Indian Institute of Science, CV Raman Avenue, Bangalore 560012, India; Medical Research Council Cognition and Brain Sciences Unit, Cambridge, UK

## Abstract

Plasma biomarkers have emerged as powerful tools to identify Alzheimer’s disease (AD) pathology and are increasingly used for diagnosis and monitoring. However, their wider differential diagnostic and prognostic value in neurodegenerative diseases other than AD remains unclear. This study tested and compared the diagnostic and prognostic performance of neurofilament light chain (NfL), p-tau217, Aβ42/40 and GFAP, in a large cohort of participants recruited from memory clinics and parkinsonism services, with survival data and neuropathology confirmation. Participants with AD and non-AD neurodegenerative diseases were recruited from memory and parkinsonism secondary healthcare services (n=646, 94 with mild cognitive impairment, 130 with Alzheimer’s dementia, 58 with Lewy-body dementia, 58 with behavioural variant frontotemporal dementia, 56 with primary progressive aphasia, 110 with progressive supranuclear palsy, 58 with corticobasal syndrome and 82 with motor neuron disease), and age-/sex-matched healthy volunteers (n=133). Out of 779 participants, 102 patients had amyloid-positivity also assessed by CSF and/or PET, and 48 patients donated their brains for neuropathological assessment. Group differences and differential performance of plasma biomarkers were analysed using non-parametric tests, ROC analyses, and Cox regression for survival prognosis. Plasma p-tau217 showed high accuracy in discriminating patients with AD *versus* controls (AUC=0.82), and patients with positive vs negative amyloid markers (AUC=0.80). However, plasma p-tau217 and p-tau231 were also elevated in patients with Lewy body dementia (*versus* controls AUC=0.73) and motor neuron disease (*versus* controls AUC=0.72). The plasma NfL/p-tau217 ratio showed better performance in differentiating patients with AD *versus* frontotemporal lobar degeneration (FTLD) pathologies at post-mortem (AUC=0.94, while p-tau217 AUC=0.82). Plasma NfL was the strongest predictor of survival (HR=1.18 [1.01-1.37], p=0.036), over and above other plasma markers and diagnoses. Our findings demonstrate the utility of plasma p-tau217 as diagnostic marker for AD pathology (noting elevation in the clinically distinct MND), and plasma NfL for prognosis across multiple neurodegenerative diseases. The NfL/p-tau217 ratio was the most sensitive and specific marker in differentiating AD from non-AD groups. We propose that the combination of plasma p-tau217 and NfL can enhance diagnostic and prognostic precision by leveraging their complementary strengths.

## Introduction

Plasma biomarkers have emerged as critical tools for diagnosing and monitoring neurodegenerative diseases. Among these, phosphorylated tau (p-tau) and neurofilament light chain (NfL) have demonstrated significant potential in identifying Alzheimer’s disease (AD) pathology and tracking disease progression respectively^1,2^. However, critical knowledge gaps affect the clinical implementation of such biomarkers into clinical practice. For example, their performance against neuropathologically confirmed aetiology, their performance in nonAD disorders, and their long term prognostic significance.

Plasma p-tau217 has translated into clinical use in many countries, because of its sensitivity to AD pathology, including strong concordance with increased amyloid plaques and tau tangles, as measured by PET imaging and neuropathology. P-tau217 levels are elevated across the continuum of AD, from preclinical stages to advanced dementia, and track disease severity, with annual changes in plasma concentrations correlated with tau PET findings ^3–10^. Plasma p-tau231 has more recently emerged as a sensitive marker for early AD pathology, showing abnormal levels even with minimal amyloid-β deposition, preceding overt plaque formation. It demonstrates high accuracy in distinguishing AD from several non-AD neurodegenerative disorders and correlates strongly with tau pathology and amyloid burden as assessed by PET imaging ^11–14^. Plasma Aβ42/40 ratios have also been widely investigated in detecting amyloid pathology. However, the ratio’s change in patients is less pronounced compared to p-tau231 or p-tau217, which may better capture early cerebral amyloid accumulation and/or tau pathology^15^. Beyond AD-specific markers, glia fibrillary acidic protein (GFAP), an astrocyte marker in brain, is a candidate for astrocyte-mediated mechanisms in plasma, correlating with disease severity, cognitive decline ^16–18^ but also amyloid pathology^17^. However, GFAP’s performance in discriminating controls from participants with mild symptoms and/or non-AD syndromes is less consistent ^19,20^. In contrast, neurofilament light chain (NfL) serves as an indicator of gross axonal damage and neurodegeneration across neurological disorders. Plasma NFL is elevated across many neurodegenerative diseases, including AD, syndromes associated with frontotemporal lobar degeneration (FTLD), motor neuron disease or amyotrophic lateral sclerosis (ALS, MND) and Lewy body dementia (LBD) ^21–26^. Unlike p-tau217, plasma NfL is not disease-specific, but it may be a valuable prognostic marker for cognitive decline and disease progression ^21–27^. Despite the progress with plasma biomarkers, few studies have examined the differential performance of plasma biomarkers across the common neurodegenerative syndromes other than AD; or tested their prognostic value. Validation studies with long term follow up, in heterogeneous clinical populations from in memory-clinic settings are urgently needed before translating biomarker research into clinical practice.

This study therefore aimed to test the diagnostic and prognostic accuracy of plasma-based biomarkers used alone or in combination (p-tau217, NfL and their ratio, p-tau231, GFAP, and Aβ1-42/Aβ1-40 ratio) in a large and clinically diverse cohort of people (n=646) recruited from memory and parkinsonism services, and healthy volunteers (n=133). Across a broad range of neurodegenerative diseases, we test their performance in differentiating by clinical syndromes, amyloid-positive vs amyloid-negative status, and pathology-confirmed cases of AD vs frontotemporal lobar degeneration (FTLD). Finally, we test their prognostic value, against long term survival.

## Materials and Methods

### Participants

Patient participants (n=646) were recruited from specialist clinics for cognitive and movement disorders at the Cambridge University Hospitals NHS Trust and collaborating regional psychiatry and neurology services. We included participants with a clinical diagnosis that met clinical consensus diagnostic criteria for amnestic mild cognitive impairment (MCI, n=94), AD-dementia (n=130) ^28,29^, Lewy-body dementia (n=58; 49 with dementia with Lewy bodies and 9 with Parkinson’s disease dementia) ^30,31^, behavioural variant Frontotemporal dementia (bvFTD; n=58); primary progressive aphasia (PPA, n=56, consisting of 31 with non-fluent variant PPA and 25 with semantic variant PPA) ^32,33^, motor neuron disease or amyotrophic lateral sclerosis (MND, n=82), probable or possible progressive supranuclear palsy (PSP, n=110, mainly Richardson’s syndrome) ^34^, and corticobasal syndrome (CBS, n=58) ^35^. We also recruited n=133 healthy controls with absence of memory symptoms and no significant neurological or systemic medical illnesses. Exclusion criteria for both patient and healthy control groups included age < 40, major concurrent psychiatric illness, other severe physical illness, or a history of other significant neurological illness.

In a sub-group of patients (n=102), presence or absence of β-amyloid (interpreted as AD-pathology) was assessed with Pittsburgh compound B tracer (PiB at a cut-off of 19 centiloids ^36^) and/or CSF Alzheimer’s biomarkers at lumbar puncture (Amyloid-beta 1-42/40 ratio < 0.065 as recommended laboratory threshold from University College London Hospitals reference laboratory ^37^). This cohort with amyloid biomarkers included conditions with either high likelihood of a significant fraction with AD as main pathology or co-pathology (LBD n=25, CBS n=19, MCI n=37). Amyloid markers were not assessed systematically for research purpose in patients with a clinical diagnosis of AD-dementia, but were available in n=21 patients with this diagnosis.

Post mortem pathology diagnosis at autopsy was available in 48 patients (post mortem diagnosis: 6 AD, 23 PSP, 7 FTLD-TDP43, 3 FTLD-Pick’s Disease, 9 corticobasal degeneration).

Participants with mental capacity gave their written informed consent to take part in the study. For those who lacked capacity, their participation followed the consultee process in accordance with national law. The research protocols were approved by the National Research Ethics Service’s East of England Cambridge Central Committee, and the UK Administration of Radioactive Substances Advisory Committee.

### Blood sample collection and processing

Blood samples were obtained by venepuncture and collected in EDTA tubes. They were centrifuged to isolate plasma, aliquoted and stored at −70/80 °C until further analyses. Plasma samples were analysed at the Clinical Neurochemistry Laboratory in Mölndal (Sweden). Plasma samples were thawed on wet ice, centrifuged at 500× g for 5 min at 4°C. Calibrators (neat) and samples (plasma: 1:4 dilution) were measured in duplicates. The plasma assays performed were the Quanterix Simoa Human Neurology 4-Plex E assay, measuring Aβ40, Aβ42, GFAP and NfL (Quanterix, Billerica, MA), p-tau231 (in-house Simoa assay) and the p-tau217 ALZpath assay, as previously described ^13,16,38^. All plasma markers are reported in pg/mL. Plasma samples were analyzed at the same time using the same batch of reagents. A four-parameter logistic curve fit data reduction method was used to generate a calibration curve. Two control samples of known concentration of the protein of interest (high-control and low-control) were included as quality control. Intra-assay coefficients of variation were below 10%.

### Statistical analysis

Analyses were performed using R (version 4.4.2). For descriptive statistics chi-square and analysis of variance tests were used to compare variables between groups. The Αβ42 and Aβ40 analytes were combined to derive the Aβ42/Αβ40 ratio, and NfL/p-tau217 ratio was calculated. Statistical analyses were performed on p-tau217, p-tau231, NfL, GFAP, Aβ42/Αβ40 ratio, and NfL/p-tau217 ratio levels. Prior to statistical analyses, outliers were excluded after being defined as values over or below 5 standard deviations from the marker-specific mean, to exclude extreme values due to methodological issues rather than pathophysiological changes (outliers: 3 p-tau217, 4 p-tau231, 9 NfL, 1 Aβ42/Αβ40, 4 NfL/ptau217).

First, we ran group comparisons with Kruskal-Wallis tests, with Dunn’s post-hoc tests, on each plasma marker separately. Classification analyses were performed using receiver operating characteristic curve (ROC) analyses to estimate the diagnostic ability of each marker, comparing each clinical diagnostic group to healthy controls. The area under the curve (AUC) was tested for each comparison.

Second, in the subgroup of patients with *in vivo* amyloid PET and/or CSF markers, we ran group comparisons on amyloid positive vs negative participants, applying a Wilcoxon signedrank test on each plasma marker separately. Classification analyses were performed using ROC curve analyses to estimate the ability of each marker to identify amyloid positivity, and the related AUCs were reported.

Third, we cross-match availability of *in vivo* markers and/or post mortem pathology confirmation, to classify participants based on the underpinning pathology: (i) AD pathology cohort: patients with a clinical diagnosis of MCI or dementia and positive amyloid PET/CSF markers, or patients with primary AD pathology at autopsy; (ii) FTLD pathology: patients with evidence of FTLD-tau (CBD, PSP, PiD) or FTLD-TDP43 as primary pathology at autopsy. This classification identified 83 patients with likely AD pathology and 42 with FTLD pathology. For each plasma marker, we ran group comparisons with Wilcoxon signedrank tests. Classification analyses were performed using ROC curve analyses to estimate the ability of each marker to differentiate AD vs FTLD. The AUC was tested for each comparison.

Fourth, we investigated the relationship between plasma markers and survival. Given the clinical heterogeneity of the cohort, we used survival data as a definitive marker of clinical progression rather than cognitive scores. Survival data were available for 528 (82%) of the patients (327 deceased and 201 still alive; 52 MCI, 60 AD, 54 LBD, 24 svPPA, 30 nfPPA, 58 bvFTD, 82 MND, 110 PSP and 58 CBS). Survival analysis used a Cox proportional hazards regression model (R function coxph), including all plasma markers (p-tau217, p-tau231, NfL, GFAP, Aβ42/Αβ40 ratio, and NfL/p-tau217 ratio), with time from blood test to death as the moderating variable of interest. Age, sex and diagnosis group were added as covariates. Plasma variables and age were centred and scaled (*scale* R function) before entering the analysis.

## Results

### Cohort characteristics

Participant demographics and clinical characteristics are described in Table 1. Patients with MCI, AD, LBD and PSP were on average older than controls, and the cohort of patients with LBD was overrepresented by males over females.

**Table 1.** Demographic and clinical characteristics of the participants included (* = significant difference between patient group and controls in post hoc analysis, p < .05).

| Group | N | Age<br>(mean (SD)) | Sex<br>(Female/Male) | PiB and<br>CSF<br>Amyloid<br>(Aβ-/Aβ+) | Pathology<br>confirmation<br>(AD/FTLD) | Survival<br>(Alive/Dead) |
| --- | --- | --- | --- | --- | --- | --- |
| Controls | 133 | 68.08 (7.75) | 63/70 | - | - | - |
| MCI | 94 | 75.46 (6.83)* | 41/53 | 2/35 | - | 39/13 |
| AD | 130 | 73.96 (9.17)* | 68/62 | 0/21 | 2/0 | 39/21 |
| LBD | 58 | 74.23 (7.15)* | 13/45* | 9/16 | - | 22/32 |
| svPPA | 25 | 68.51 (4.77) | 9 /16 | - | 0/4 | 10/14 |
| nfPPA | 31 | 71.74 (7.22) | 15/16 | - | 2/3 | 12/18 |
| bvFTD | 58 | 64.30 (8.60) | 21/37 | - | 0/6 | 21/37 |
| MND | 82 | 66.63 (10.21) | 37/45 | - | - | 19/63 |
| PSP | 110 | 71.62 (7.37)* | 50/60 | - | 0/18 | 21/89 |
| CBS | 58 | 70.50 (6.33) | 35/23 | 12/7 | 2/11 | 18/40 |
| Group<br>Comparisons | - | F(9)=16.2,<br>p<0.001 | X2(9)=23.4,<br>p=0.005 | - | - | - |

### Plasma markers across clinical diagnoses

Group comparisons on plasma markers confirmed diagnostic differences, with KruskalWallis tests being significant across all six tested markers. Specifically, p-tau217 levels (Kruskal-Wallis test (9)=158, p<0.001) were elevated in patients with clinical diagnoses of MCI, AD, LBD, MND and CBS compared to healthy controls (Figure 1A). The ROC curve analysis for p-tau217 across the nine diagnostic groups as compared to controls reveals distinct patterns in sensitivity and specificity (Figure 1B). Patients with AD and MCI exhibit the highest AUC values of 0.82 and 0.76 respectively, indicating great discriminatory ability. In contrast, groups of patients with PSP and bvFTD showed the lowest AUC values of 0.53 and 0.55, respectively, indicating low discrimination, with LBD being intermediate at 0.73.

**Figure 1.**
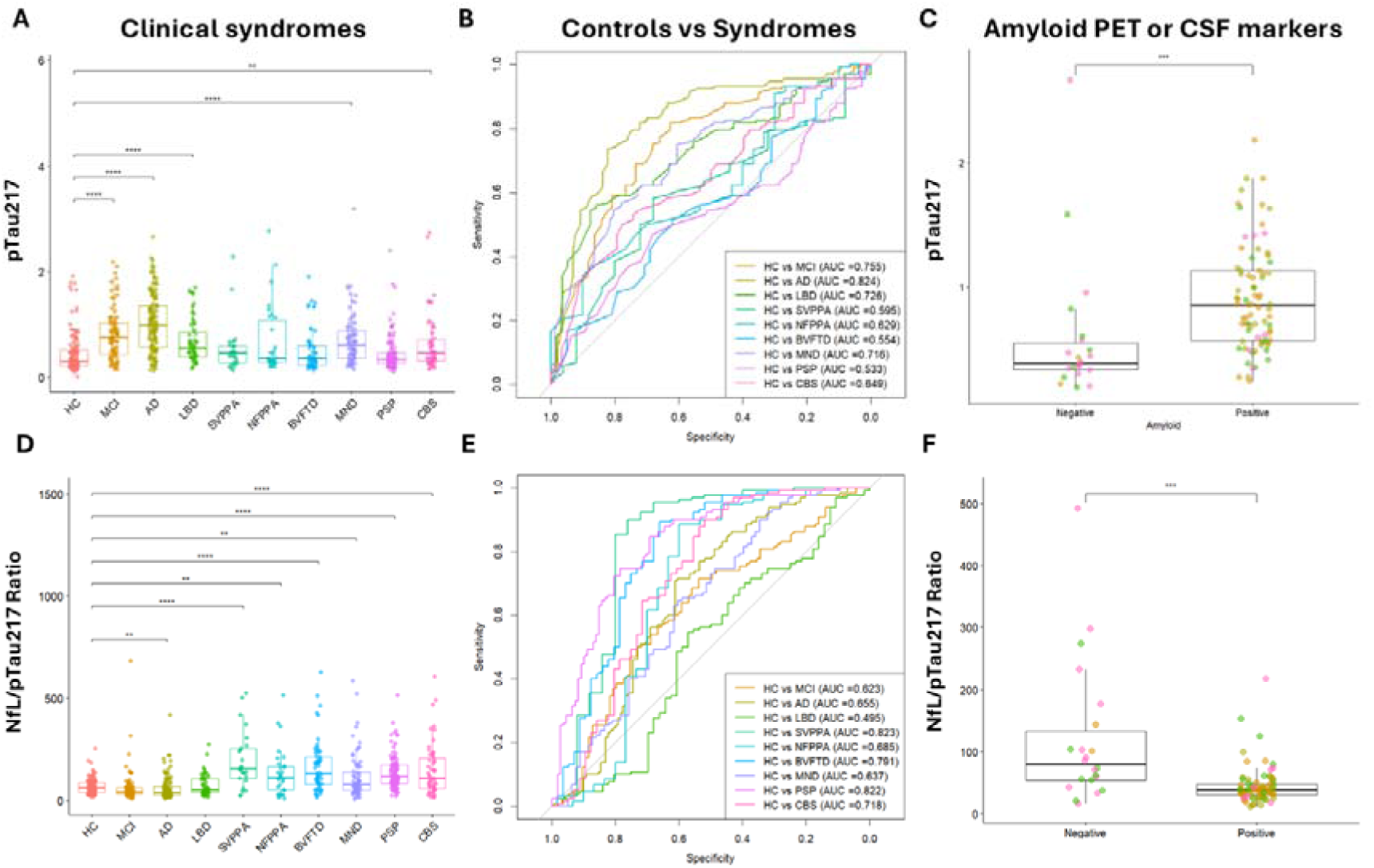
Plasma p-tau217 levels and NfL/p-tau217 ratio differentiate controls from patients with different diagnoses, and amyloid positive vs negative patients.

The NfL/p-tau217 ratio showed the highest group effects in non-AD cohorts (Kruskal-Wallis test (9)=197, p<0.001), when comparing clinical syndromes with controls (Figure 1D). The ROC curve analysis and AUC values indicated highest sensitivity and specificity in differentiating controls from patients with PSP (AUC=0.82) and svPPA (AUC=0.82), followed by bvFTD (AUC=0.79) and CBS (AUC=0.72) (Figure 1E). See Supplementary Figure 1 for distribution of patients with different clinical diagnoses across quartiles of NfL/p-tau217 ratio: an increasing proportion of patients with non-AD clinical syndromes was observed from quartile 1 to quartile 4. The NfL/p-tau217 ratio had a better performance in discriminating group of patients with AD vs patients with SVPPA (AUC=0.855), NFPPA (AUC=0.728), bvFTD (AUC=0.835), MND (AUC=0.733), PSP (AUC=0.851) and CBS (AUC=0.779) Supplementary Figure 2B), than p-tau217 alone (Supplementary Figure 2A). The NfL/p-tau217 ratio showed poor performance in differentiating controls from patients with LBD (AUC=0.495), who were more equally distributed across the ratio quartiles (Supplementary Figure 1). See Supplementary Figure 2 for all AUCs from pairwise comparisons on p-tau217 (2C) and NfL/p-tau217 ratio (2D).

Similarly to p-tau217, p-tau231 levels p-tau231 (Kruskal-Wallis test (9)=130, p<0.001) were significantly elevated in patients with clinical diagnoses of MCI, AD, LBD, and MND as compared to controls, but not CBS (Figure 2A). The MND cohort showed the highest AUC values of 0.81. Plasma levels of NfL (Kruskal-Wallis test (9)=179, p<0.001) were elevated in all diagnostic groups, while GFAP levels (Kruskal-Wallis test(9)=123, p<0.001) in all diagnostic group but not in patients with svPPA or bvFTD (Figure 2B and 2C). In contrast, group comparison analyses on Aβ42/Aβ40 ratio (Kruskal-Wallis test(9)=13.3, p=0.147) did not identify any differences between patients and controls, and showed low AUC values for all diagnostic groups (Figure 2D).

**Figure 2.**
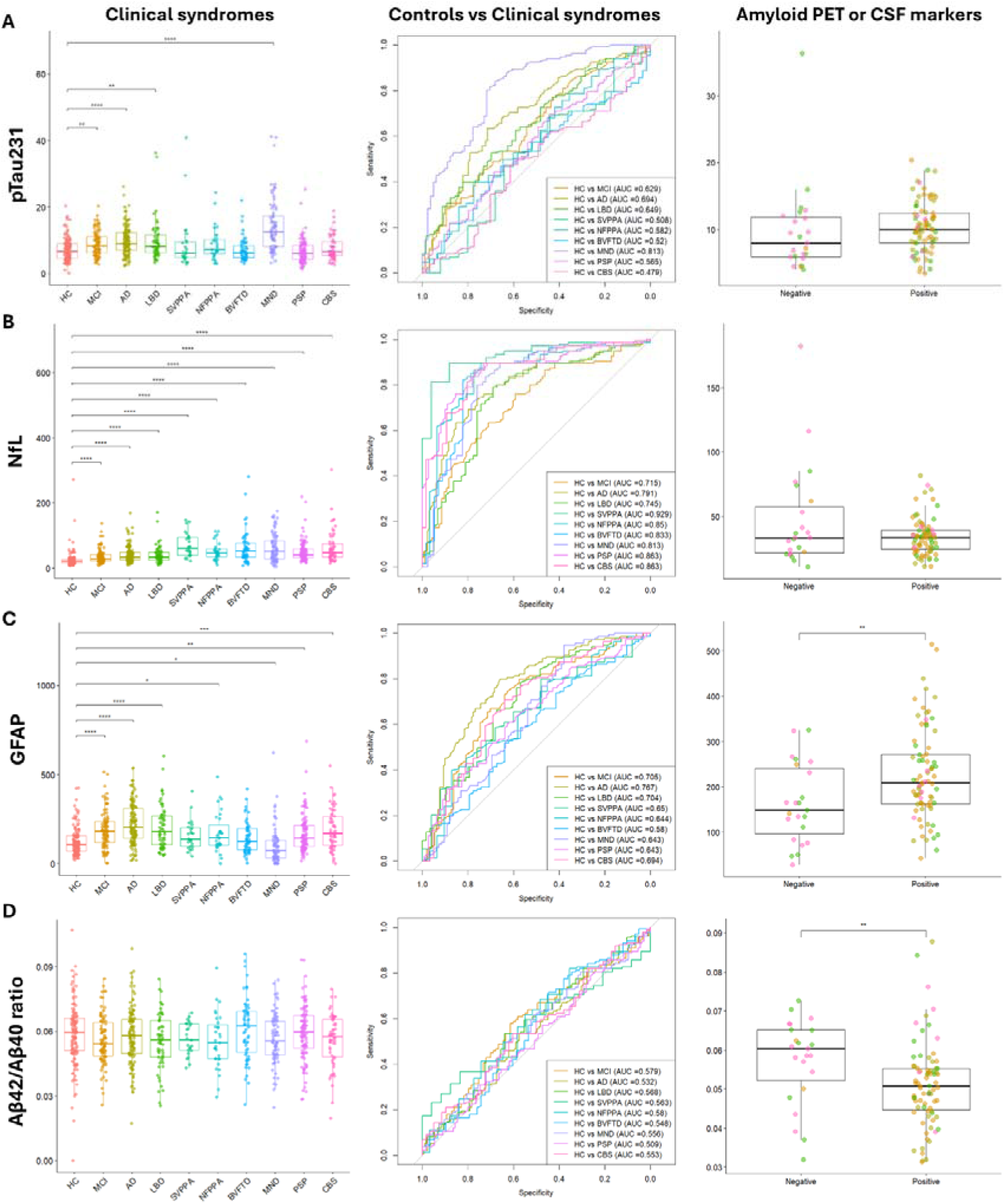
Discriminatory performance of p-tau231, NfL, GFAP and Aβ42/Aβ40 ratio for controls vs different diagnoses, and amyloid positive vs negative patients.

### Plasma markers differentiate amyloid-positive vs amyloidnegative patients

Next, in patients with *in vivo* amyloid PET and/or CSF markers, we ran group comparisons comparing amyloid-positive with amyloid-negative participants. Wilcoxon tests on plasma markers identified statistically higher plasma levels of p-tau217 (W=347.5, p<0.001, Figure 1C) and GFAP (W=568, p=0.0065, Figure 2C) and lower values of the NfL/p-tau217 (W=1377, p<0.001, Figure 1F) and Aβ42/Aβ40 (W=1284, p=0.0027, Figure 2D) ratios in amyloid-positive patients. In contrast, plasma p-tau231 (W=691, p=0.08, Figure 2A) and NfL (W=965.5, p=0.651, Figure 2B) were not different between the two groups.

### Plasma markers in AD and FTLD confirmed pathology

Based on the availability of *in vivo* markers and/or post-mortem pathology confirmation, we identified a sub-group of patients with AD (n=83) or FTLD pathology (n=42). Wilcoxon tests on plasma markers comparing AD vs FTLD cohorts identified statistically differences on all markers (Supplementary Figure 3). The ROC curve analyses comparing each marker in the two pathology cohorts with healthy controls showed distinct patterns in sensitivity and specificity (Figure 3). Plasma p-tau217 (AUC=0.86) and GFAP (AUC=0.81) levels showed great discriminatory ability in comparing controls with patients with AD pathology. NfL and NfL/p-tau217 ratio showed the highest AUC values in comparing patients with FTLD pathology and controls (AUC=0.91 and AUC=0.94, respectively).

**Figure 3.**
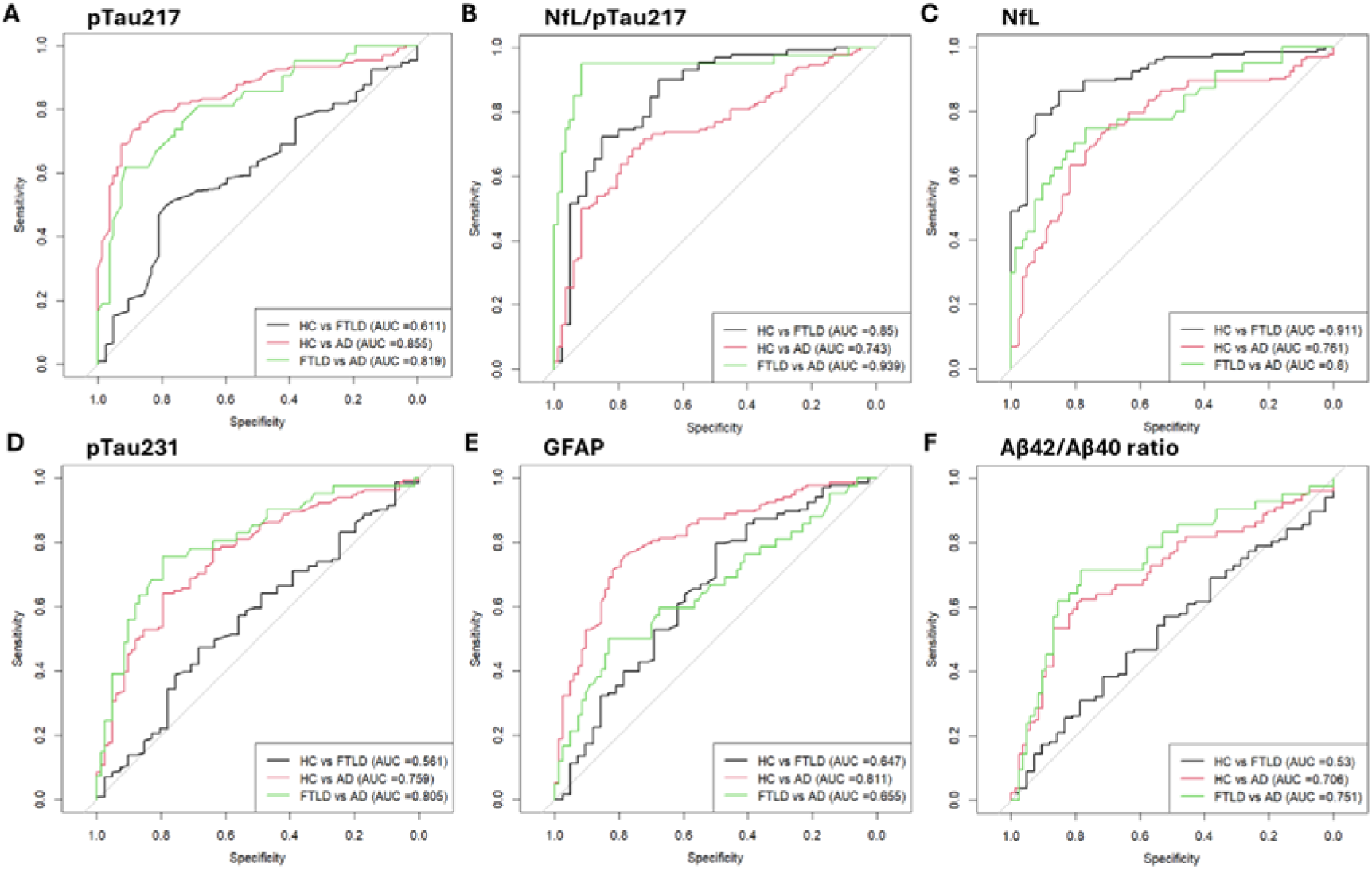
Classification performance for pathology detection of each plasma marker, expressed as area under the curve (AUC).

### Comparisons of plasma markers in Aβ+/Aβ- and AD/FTLD pathology groups

Comparing the plasma markers on their ability to discriminate amyloid-positive vs amyloid-# negative patients (Figure 4, left panel), the ROC curve analyses and AUCs indicated that the p-tau217 had better overall discriminatory ability (AUC=0.80), with sensitivity of 0.86 and specificity of 0.73, followed by NfL/p-tau217 ratio (AUC=0.79; sensitivity: 0.79 and specificity: 0.77) and Aβ42/Aβ40 ratio (AUC=0.71; sensitivity: 0.81 and specificity: 0.70). Plasma GFAP (AUC=0.69; sensitivity: 0.81 and specificity: 0.30), p-tau231 (AUC=0.62; sensitivity: 0.76 and specificity: 0.52) and NfL (AUC=0.47; sensitivity: 0.81 and specificity: 0.30) showed lower discriminatory ability. This analysis identified 0.50 pg/mL as the threshold for plasma p-tau217 in the comparison of amyloid-positive vs amyloid-negative groups.

**Figure 4.**
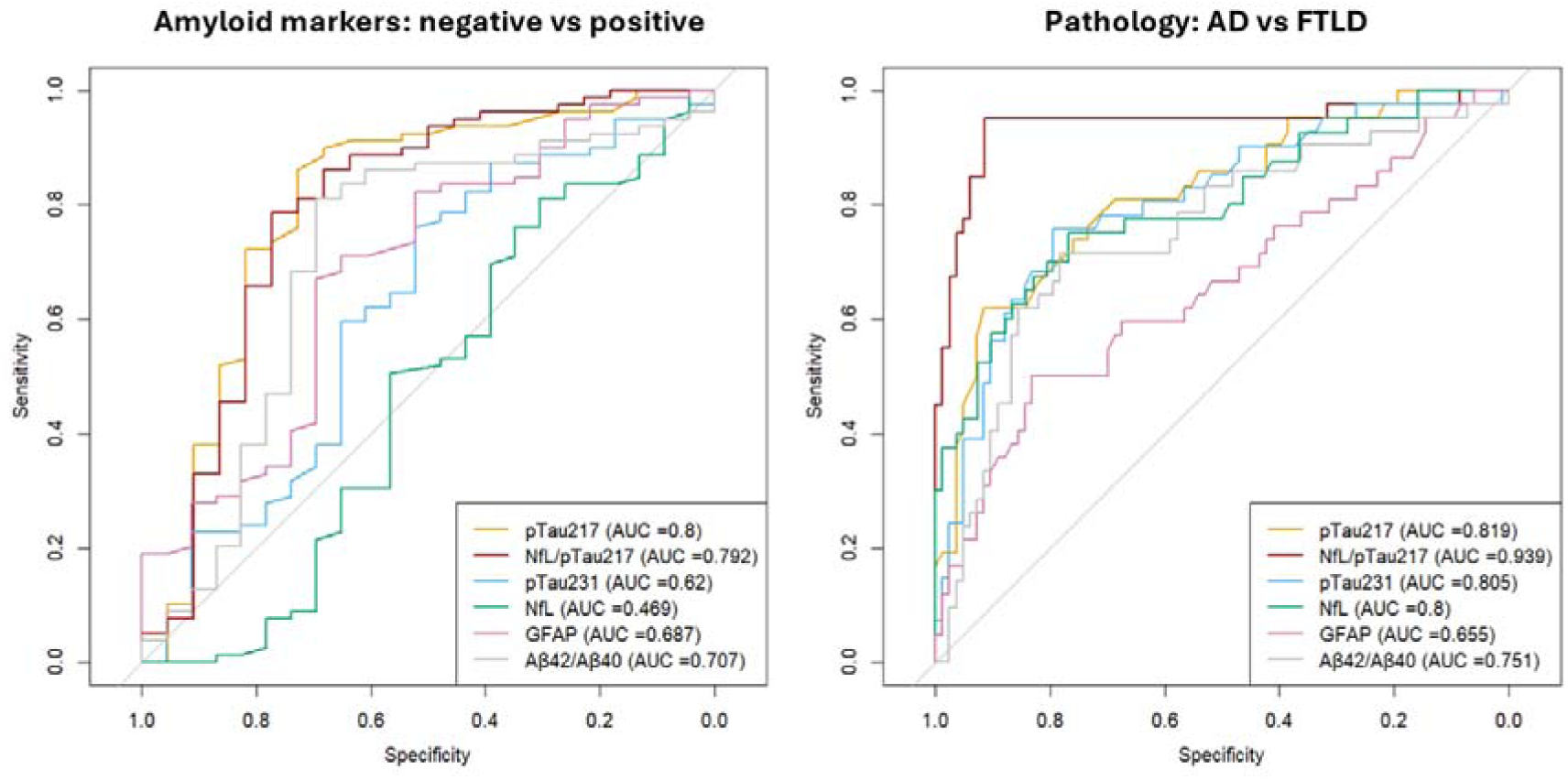
Direct comparison of plasma marker performance in discriminating Aβ+ vs. Aβ− participants and Alzheimer’s disease (AD) vs. frontotemporal lobar degeneration (FTLD) cohorts.

In head-to-head comparisons of the markers’ ability to discriminate AD and FTLD pathology (Figure 4, right panel), the ROC curve analyses and AUCs indicated that the NfL/p-tau217 ratio had the best overall discriminatory ability (AUC=0.94), with sensitivity of 0.95 and specificity of 0.92, followed by p-tau217 (AUC=0.82; sensitivity: 0.62 and specificity: 0.92), p-tau231 (AUC=0.81; sensitivity: 0.76 and specificity: 0.80), NfL (AUC=0.80; sensitivity: 0.75 and specificity: 0.77), and Aβ42/Aβ40 ratio (AUC=0.75; sensitivity: 0.71 and specificity: 0.78). Plasma GFAP showed the lowest AUC value (AUC=0.66), with sensitivity of 0.50 and specificity of 0.83. This analysis identified 0.48 pg/mL as the threshold for plasma p-tau217 in the comparison of amyloid-positive vs amyloid-negative groups.

### Threshold effects

Ashton et al. proposed a two-threshold approach for amyloid positivity based on plasma ptau217 ^39^. Accordingly, we considered p-tau217 >0.63 pg/mL for amyloid-positivity and high likelihood of AD pathology, and p-tau217 levels 0.4-0.63 pg/mL being an “intermediate” zone, while p-tau217 <0.4 pg/mL indicating amyloid negativity. Patients with MCI and AD mostly met the criteria for amyloid positivity with p-tau217 levels >0.63 pg/mL, as well as a large number of patients with a clinical diagnosis of LBD and MND. In contrast, most patients with FTLD-related syndromes and FTLD-confirmed pathology had lower p-tau217 levels. See Supplementary Figure 4.

### Plasma NfL predicts survival, over and above other plasma markers and diagnosis

We tested the prognostic value of the plasma markers (in patients only) applying Cox proportional hazards regression with days from blood test to death or census as outcome, plasma levels of p-tau217, NfL/p-tau217 ratio, p-tau231, NfL, GFAP, Aβ42/Αβ40 ratio as predictors of interest, and age, sex, disease groups as covariates. The model identified a significant predictive effect of NfL levels (HR=1.18 [1.01-1.37], p=0.036), age at blood test (HR=1.24 [1.09-1.42], p=0.001), and diagnoses, over and above the other plasma markers and sex (Figure 5, left panel; and Supplementary Table 1). High NfL, older age, all diagnosis as compared to MCI were associated with poor survival. To illustrate the prognostic effect of NfL (Figure 5, right panel), we plotted Kaplan-Meier curves dividing patients with high and low values of NfL, as separated by the median.

**Figure 5.**
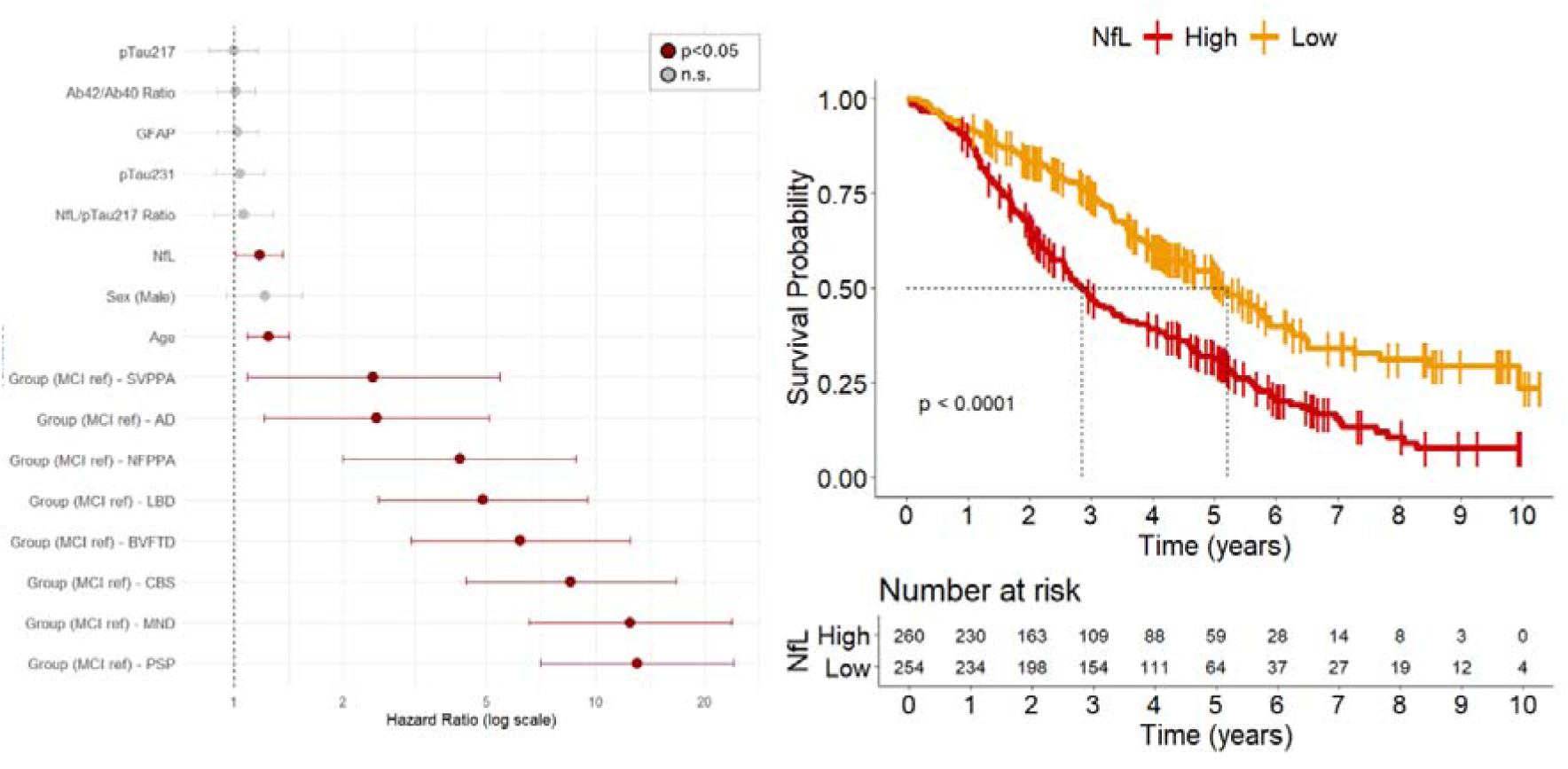
Prognostic value of biomarkers against mortality. Left panel: Hazard ratio values of the survival model with all plasma markers of interest, age, sex and diagnosis as predictors. Right panel: Kaplan–Meier survival curve of NfL: patients were separated into two groups based on the median split of NfL plasma levels at baseline.

We used Cox proportional hazards regression with the predictors significantly associated with survival rates in the full survival model, including plasma NfL levels, age and diagnostic group, and their interactions (NfL*group*age). The model identified a significant interaction effect between age and diagnosis on survival (χ²(8)=19.7, p=0.01), but not of NfL*age (χ²(8)=0.4, p=0.55) or NfL*diagnosis (χ²(8)=13.8, p=0.09).

## Discussion

This study demonstrates the complementary information conveyed by plasma biomarkers, in the context of multiple neurodegenerative disorders, for differential diagnosis and prognosis. Plasma p-tau217 was confirmed as a selective diagnostic marker for AD pathology (against PET and post mortem gold standard), except for its marked elevation in LBD, MND/ALS, and FTLD-related syndromes. Plasma NfL/p-tau217 ratio was the most sensitive and specific marker in differentiating patients with non-AD syndromes from patients with AD and controls, and between patients with AD vs FTLD pathology. In contrast, baseline plasma NfL indicates prognosis across multiple neurodegenerative diseases. The elevation of p-tau217 and p-tau231 in people with MND is unlikely to be due to diagnostic error, due to the high and distinctive clinic-pathological correlations.

Different plasma p-tau epitopes reflect different pathological processes related to AD, with ptau231 being associated with the earliest amyloid-β increases and p-tau217 with both amyloid and tau pathology accumulation ^2,12,13^. Our findings align with previous evidence on plasma p-tau biomarkers showing higher diagnostic performance and larger effect sizes than Aβ42/40 assays in detecting AD pathology, with p-tau217 assays consistently outperforming the others (including p-tau181 and p-tau231) ^3,4,14,15,40,41^. In our clinic-based recruited cohort, we confirmed that p-tau217 could identify amyloid-β pathology in individuals with dementia and confirmed AD pathology by autopsy and/or amyloid CSF/PET biomarkers, in line with previous evidence in large cohorts ^2,39,41,42^. Across several studies, plasma p-tau217 assays have been found strongly correlated with p-tau217 CSF levels, amyloid and tau PET, and with cognitive performance across the AD spectrum ^3–10^. Plasma p-tau217 is also a good prognostic marker for predicting future cognitive decline in cognitively unimpaired individuals ^40,43,44^, with similar performance to tau-PET ^45^, suggesting its potential as screening tool in preclinical AD trials. Towards further validation for the use of plasma ptau217 at individual level for clinical trials and patient screening, the thresholds identified in our analyses for plasma p-tau217 comparing amyloid positive vs negative patients (0.50 pg/mL) and AD vs FTLD pathology (0.48 pg/mL) were in the same range of previously described thresholds by Ashton et al. ^39^, with p-tau217 >0.63 pg/mL indicating amyloid-positivity and high likelihood of AD pathology, and p-tau217 levels 0.4-0.63 pg/mL being an “intermediate” zone.

Plasma p-tau epitopes have been shown to be a useful tool to capture AD as primary or copathology in multiple neurodegenerative syndromes, including syndromes associated with FTLD ^46–48^ and LBD ^49–51^. In our data, elevated plasma p-tau217 and p-tau231 levels were identified in clinical syndromes often associated with AD pathology or co-pathology, including MCI, clinical Alzheimer’s dementia, LBD and CBS. However, we also found increased p-tau217 and p-tau231 in most of the patients with MND. This aligns with and expands on a recent study showing elevated p-tau181 and p-tau217 serum levels in patients with amyotrophic lateral sclerosis (ALS) compared to controls, and the presence of p-tau181 and p-tau217 in muscle biopsies from ALS cases: with increased p-tau reactivity in atrophic muscle fibres ^52^. The elevation of plasma p-tau217 and p-tau231 in patients with MND in our cohort may then reflect the release of p-tau from denervated muscle fibres. A recent study showed differential results in MND, based on the p-tau217 assay, but the origins of this pTau217 signal in MND may be distinct from AD. Specifically, ALZpath p-tau217 assay targets both low-molecular-weight and high-molecular-weight tau, with the latter being present in the peripheral nervous system hence elevated in MND. In other assays, targeting low-molecular-weight tau only, this increase is not observed, and the assay has greater AD specificity ^53^. Further studies are needed to clarify the presence and the role of peripheral tau pathology in MND and neuromuscular diseases, and the influence of high- versus low-molecular weight assays on the identification of p-tau217 in ALS/MND.

Blood levels of NfL are a proxy marker of the rate of neuroaxonal degeneration and this may underlie the prognostic value across neurodegenerative disorders. Serum and plasma NfL levels have been increasingly implemented as stratification tools, endpoints or surrogate markers in clinical trials. NfL is also in use in several countries to confirm neurodegeneration as an underlying cause of symptoms in primary care or memory clinics ^21–26^. In our cohort, increased levels of plasma NfL were identified across all patient groups, with the most elevated levels in patients with FTLD-associated syndromes including MND. Plasma NfL also showed the strongest prognostic performance in predicting survival rates across all diagnostic groups. This effect in AD and DLB goes beyond previous evidence for the association of high plasma or serum NfL levels with shorter survival, in patients with FTLD-associated syndromes ^47,54–58^ and MND ^59,60^.

Combining biomarkers like p-tau217 and NfL can increase their performance for diagnostic precision, supporting clinical decision-making, and future applications in personalized medicine ^61^. Low p-tau217 levels paired with low NfL has been associated mostly with nonneurological diagnoses (79%), while and high p-tau217 paired with low NfL indicated AD pathology at any stage (84%) ^49^. Here, we examined the utility of combining the two markers as ratio of plasma NfL to p-tau217 levels. Clinically, the NfL/p-tau217 ratio showed better accuracy than p-tau217 in discriminating patients with AD from patients with FTLD-associated clinical syndromes. Looking at pathology, lower NfL/p-tau217 values were found in amyloid-positive patients as compared to amyloid-negative participants, and the NfL/ptau217 ratio was the most accurate measure in differentiating AD vs FTLD-pathology confirmed cohorts. This result indicates that the combination of p-Tau217 and NfL holds greater potential for clinical application.

This study has limitations. First, patients were recruited from memory and parkinsonism clinics based on their clinical diagnosis, and pathology confirmation was available only in a sub-cohort included in the study. Misdiagnosis and/or co-pathologies may occur. This risk is mitigated by the consistency of results in the subgroup with neuropathological confirmation. Second, our cohort was predominantly white/Caucasian reflecting the ethnicity distribution of the over 65-year-old population in the UK (94% ‘white’ in the 2021 national census). The performance of blood-based biomarkers may vary in other populations. Third, we have not accounted for factors that may impact on the quantification of the tested plasma markers, such as kidney dysfunction ^62^, comorbid medical conditions ^63^, concomitant medication and variation with time of day ^64^ (our samples were acquired between 10am and 5pm). Further studies are needed to test the generalisation of our results to other cohorts and to identify methodological, environmental and genetic influences.

In conclusion, our findings provide evidence that plasma markers p-tau217, p-tau231 and NfL are useful tools for diagnosing and managing neurodegenerative diseases. In particular, p-tau217 is highly effective in identifying AD pathology across multiple syndromes, while NfL indicated the aggressiveness of neurodegeneration and future clinical decline. Plasma NfL/p-tau217 ratio was the most sensitive and specific marker in differentiating patients with non-AD syndromes from patients with AD and controls, and between patients with AD vs FTLD pathology. The combination of plasma p-tau217 and NfL can enhance clinical practice and trials by leveraging their complementary strengths. Using them together can improve diagnostic accuracy, empower clinical decision-making, patient stratification and personalised medicine in neurodegenerative diseases.

## Data Availability

Anonymized processed data can be shared upon request to the corresponding author. Access to raw data may also be requested but is likely to require a data transfer agreement with restrictions required to comply with participant consent and data protection regulations.

## Acknowledgments

We thank our participant volunteers for their participation in this study, thank the National Institute for Health Research (NIHR) Cambridge BioResource centre staff, and the research nurses for their contribution, and the East Anglia Dementias and Neurodegenerative Diseases Research Network (DeNDRoN) for help with subject recruitment.

For the purpose of open access, the authors have applied a Creative Commons Attribution (CC BY) license to any Author Accepted Manuscript version arising from this submission. This work is licensed under a Creative Commons Attribution 4.0 International License.

## Supplementary material

Supplementary material is available at *Brain* online

## Abbreviations

(NfL): neurofilament light chain
(p-tau): phosphorylated tau
(GFAP): glia fibrillary acidic protein
(FTLD): frontotemporal lobar degeneration
(ALS, MND): motor neuron disease or amyotrophic lateral sclerosis
(LBD): Lewy body dementia.

