## Supplementary Material for "Clinicopathological and prognostic correlations of plasma biomarkers in neurodegenerative diseases"

**Author affiliations:**

*Supplementary table 1. Survival model with plasma markers, age, sex and diagnosis. P values < 0.05 of predictors in the final model are highlighted in yellow.*

|  | **HR** | **SE** | **Lower 95%** | **Upper 95%** | **z** | **Pr(>\|z\|)** |
| --- | --- | --- | --- | --- | --- | --- |
| **pTau217 (std)** | 0.997 | 0.080 | 0.852 | 1.165 | -0.041 | 0.967 |
| **pTau231 (std)** | 1.037 | 0.079 | 0.888 | 1.212 | 0.459 | 0.646 |
| **NfL (std)** | 1.174 | 0.077 | 1.011 | 1.365 | 2.097 | 0.036 |
| **GFAP (std)** | 1.022 | 0.068 | 0.895 | 1.167 | 0.319 | 0.750 |
| **Aβ42/40 Ratio (std)** | 1.012 | 0.062 | 0.897 | 1.143 | 0.199 | 0.842 |
| **NfL/pTau217 Ratio (std)** | 1.062 | 0.095 | 0.882 | 1.280 | 0.638 | 0.523 |
| **Age (std)** | 1.243 | 0.067 | 1.090 | 1.419 | 3.235 | 0.001 |
| **Sex (Male)** | 1.214 | 0.122 | 0.956 | 1.543 | 1.591 | 0.112 |
| **Group (MCI ref) - AD** | 2.478 | 0.366 | 1.209 | 5.078 | 2.478 | 0.013 |
| **Group (MCI ref) - LBD** | 4.887 | 0.340 | 2.510 | 9.516 | 4.666 | 0.000 |
| **Group (MCI ref) - SVPPA** | 2.430 | 0.410 | 1.089 | 5.422 | 2.167 | 0.030 |
| **Group (MCI ref) - NFPPA** | 4.206 | 0.378 | 2.005 | 8.820 | 3.802 | 0.000 |
| **Group (MCI ref) - BVFTD** | 6.198 | 0.355 | 3.092 | 12.423 | 5.142 | 0.000 |
| **Group (MCI ref) - MND** | 12.484 | 0.329 | 6.557 | 23.770 | 7.684 | 0.000 |
| **Group (MCI ref) - PSP** | 13.041 | 0.313 | 7.055 | 24.107 | 8.192 | 0.000 |
| **Group (MCI ref) - CBS** | 8.550 | 0.341 | 4.385 | 16.673 | 6.298 | 0.000 |

*Supplementary Figure 1. Proportion of patients with different clinical diagnoses across NfL/p-tau217 quartiles.*

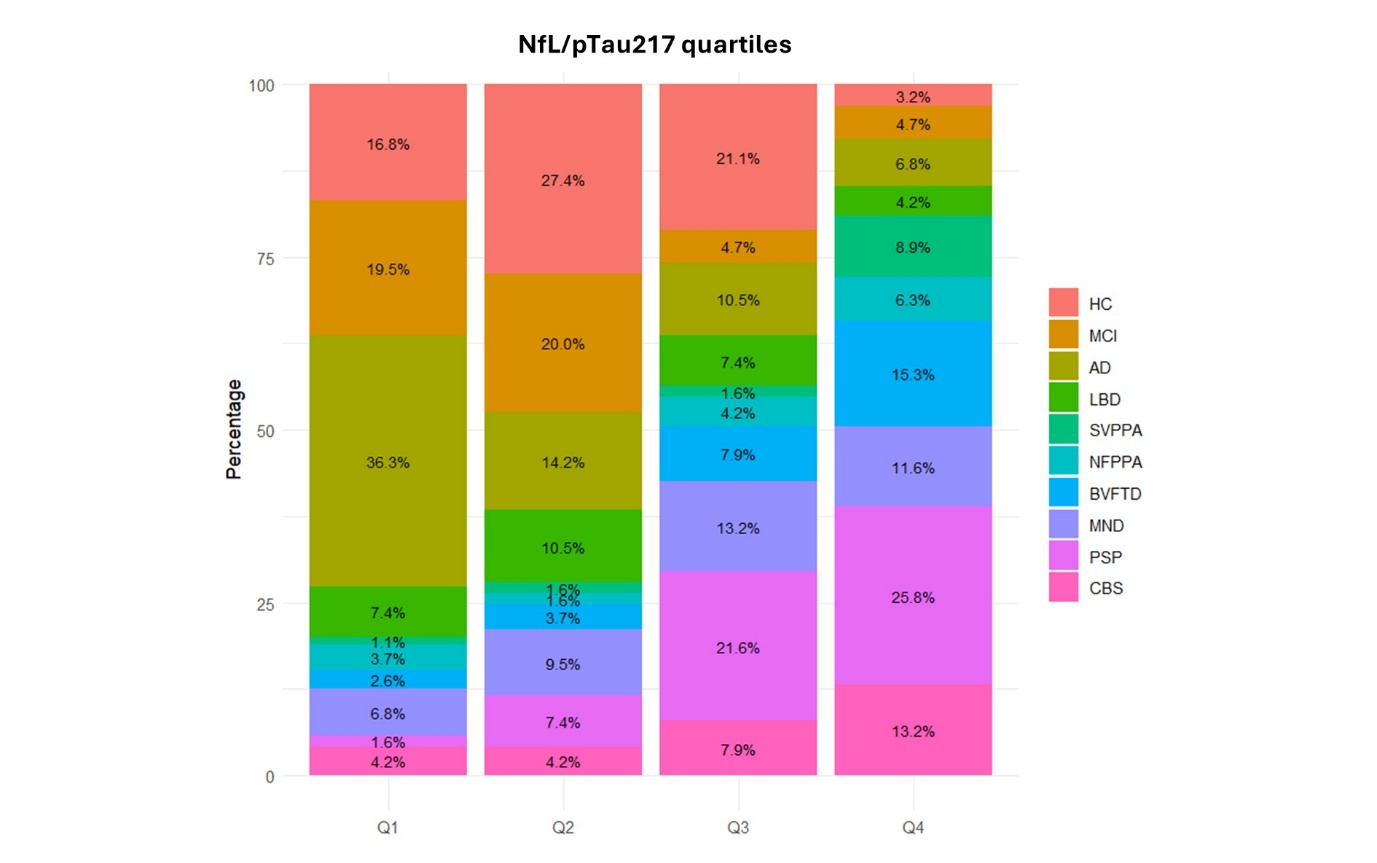

*Supplementary Figure 2. Direct comparisons of p-tau217 and NfL/p-tau217 ratio’s performance in discriminating patients with a diagnosis of AD vs other groups (A and B respectively), and across all pairwise group comparisons (C and D).*

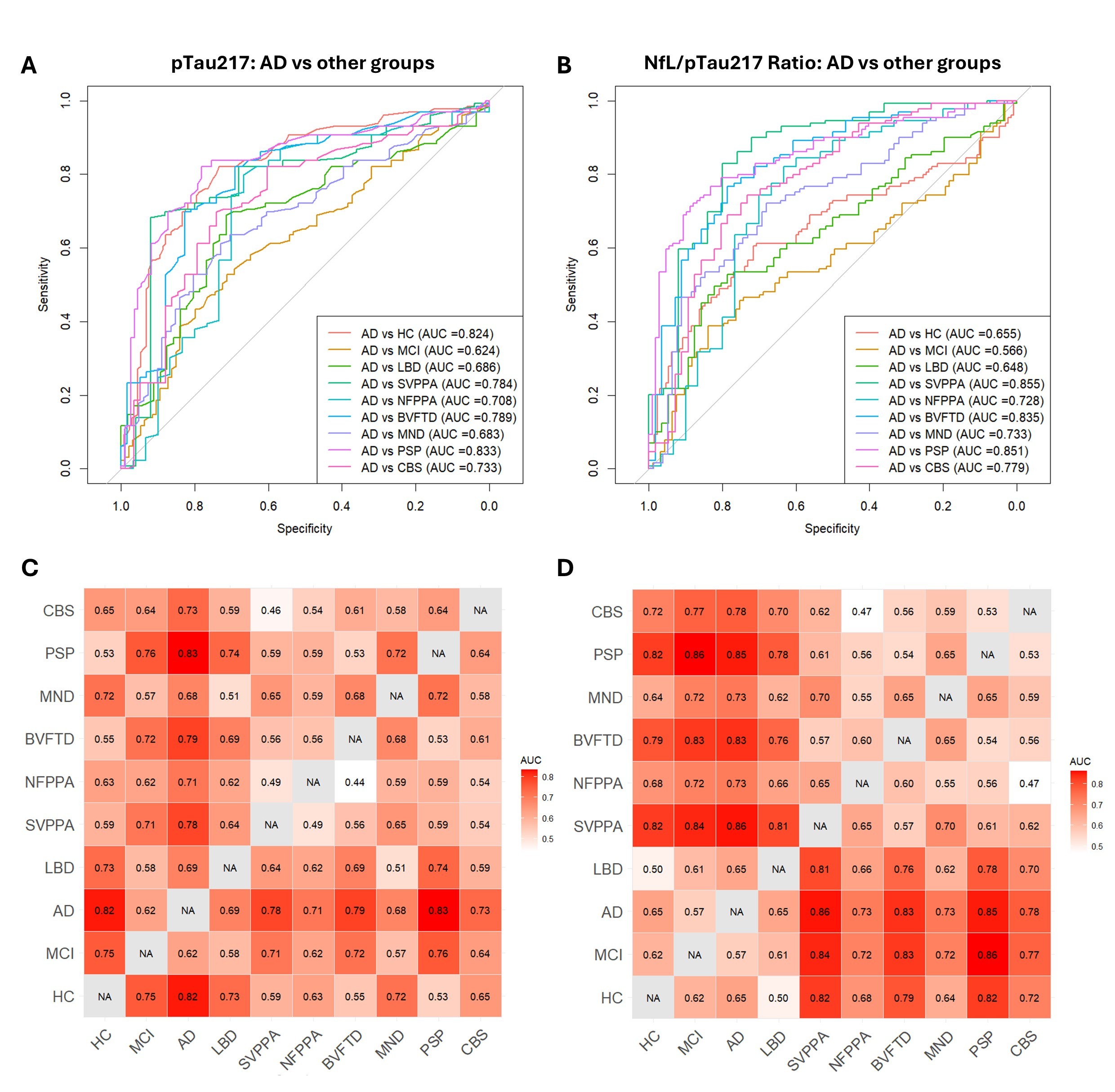

*Supplementary Figure 3. Comparisons on plasma markers in AD and FTLD cohorts with confirmed pathology.*

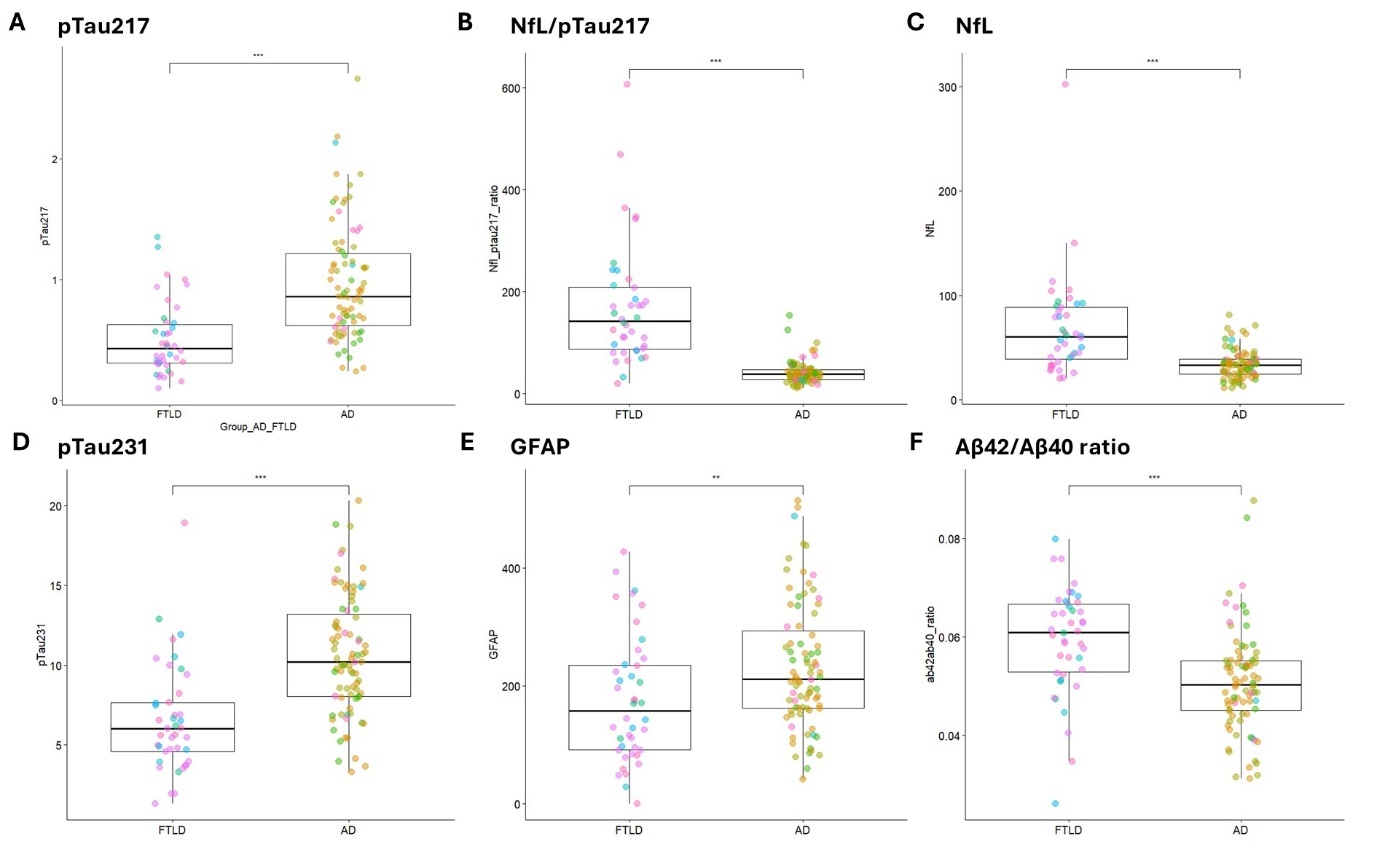

*Supplementary Figure 4. Plasma p-tau217 performance across clinical syndromes and pathological groups.*

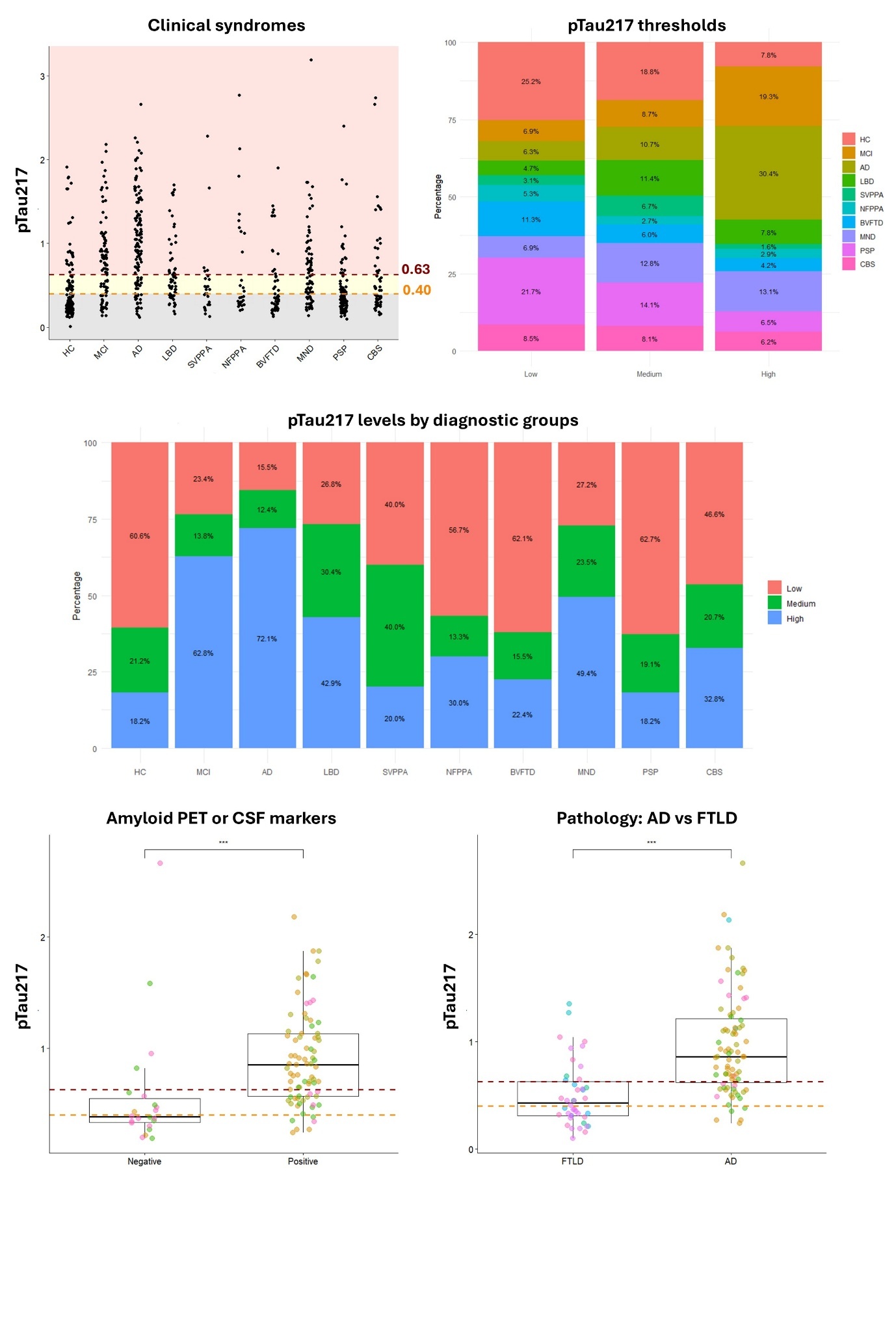
